# A prospective controled randomized multicenter study to evaluate severity of compensatory sweating after one-stage bilateral thoracic sympathectomy versus unilateral thoracic sympathectomy in the dominant side

**DOI:** 10.64898/2026.02.18.26346562

**Authors:** Miguel Lia Tedde, Niura Noro Hamilton, Nelson Wolosker, Marina Borri Wolosker, Wolfgang W Schmidt Aguiar, Hylas Paiva da Costa Ferreira, Fernando Luiz Westphal, Alexandre M Rodrigues Lima, Humberto A de Oliveira, Sergio Tadeu L F Pereira, Fabio de Oliviera Riuto, Guilherme C Resende, Marina Maria Krum Brenner, Daniel de Oliveira Bonomi, Caroline E Brero Valero, Paulo M Pego-Fernandes

## Abstract

2

**OBJECTIVE:** To compare, in a Brazilian population, the clinical efficacy and quality-of-life (QoL) impact of one-stage bilateral thoracic sympathectomy (BTS) versus unilateral sympathectomy on the dominant side (UniS), with additional analysis of patients who later underwent contralateral surgery (two-stage bilateral, 2stS).

**METHODS:** Prospective, randomized, controlled, multicenter trial (11 centers) including 163 adults with primary palmar hyperhidrosis. Participants were randomized 1:1 to BTS or UniS. From 6 months onward, UniS patients could elect contralateral sympathectomy (2stS). Sweating severity was assessed using the Hyperhidrosis Disease Severity Scale (HDSS) across 18 anatomical sites at each visit. Compensatory sweating (CS) was defined as new sweating in previously unaffected areas (preoperative HDSS = 1) and graded by the magnitude of HDSS increase. QoL was measured with two complementary validated instruments: HidroQOL and the Horn questionnaire.

**RESULTS:** Baseline characteristics were similar between groups, with most participants presenting severe preoperative disease. Improvement in the operated (dominant) hand was comparable after BTS and UniS, whereas control of the non-operated hand favored BTS. In the UniS group, spontaneous contralateral improvement occurred in approximately one-seventh of untreated hands. The proportion of patients without CS was similar in both groups (∼25%), but severe CS was more frequent after BTS (40.4% vs 21.0%, p = 0.0344). QoL improved in both groups, with larger and more sustained reductions in Horn and HidroQOL scores after BTS (p < 0.001). In the 2stS subgroup, contralateral surgery produced a consistent HDSS decrease and marked QoL improvement, with predominantly mild additional CS.

**CONCLUSIONS:** BTS provides more complete symptom control and greater QoL improvement, but at the cost of more severe CS. UniS offers excellent control on the treated side, may reduce severe CS, and supports a staged strategy in which some patients avoid a second procedure (requested by 22.5% in this study); when needed, contralateral completion tends to restore additional clinical and QoL gains.

## 3. Introduction

Hyperhidrosis (HH) is a common clinical condition that affects approximately 2,8% of the general population.^1^ It is defined as a somatic disorder characterized by excessive and uncontrolled sweating localized to specific body regions, occurring beyond what is physiologically necessary for temperature regulation.^2^ Among its various presentations, palmar hyperhidrosis (PH) is most frequent and clinically significant, as it can interfere with emotional well-being, social interactions, and professional performance.^3^ Patients usually report embarrassment, engage in avoidance behaviors, and experience occupational limitations related to hand moisture.^4^ This condition is frequently associated with plantar and axillary hyperhidrosis, forming a spectrum of hyperhidrotic disorders.^5^ Although HH is considered benign, its psychosocial burden can be substantial, sometimes leading to anxiety, low self-esteem, and social withdrawal.^6^

The most widely accepted and effective surgical treatment for PH is bilateral video-assisted thoracic sympathectomy (BTS), with both sides performed during the same anaesthetic session.^7, 8^ This minimally invasive technique offers excellent long-term symptom relief and allows a quick recovery, significantly improving patients’ quality of life (QoL).^9^

Although BTS is considered a highly safe and effective procedure, it is not entirely free from complications.^10, 11^ The most common adverse event is compensatory sweating (CS), which predominantly affects the trunk, abdomen, and back, and may occur in up to 90% of patients. In most cases, the intensity is mild and does not lead to significant functional impairment.^12^ However, despite being relatively benign for many patients, CS remains a key concern due to its potential to compromise satisfaction with surgical outcomes.^13^ The exact pathophysiological mechanisms underlying CS remain incompletely elucidated. It is believed to result from an imbalance in thermoregulation following sympathetic denervation, resulting in excessive sweating in previously unaffected areas as a compensatory response.^14^

Several variables appear to influence both the incidence and the severity of CS, including the extent and level of sympathetic chain interruption, individual differences in autonomic regulation, and environmental or psychological triggers.^15^

Additionally, patient-specific characteristics, such as baseline sweating intensity, body mass index, and environmental conditions, may further modulate symptom presentation.^16^

Understanding these determinants is essential to improve patient selection, develop preventive strategies, and refine surgical techniques to reduce the incidence of CS—ultimately enhancing overall postoperative satisfaction and QoL.

To reduce the incidence of CS, some authors have compared two different approaches: Unilateral sympathectomy (UniS) and a two-stage sympathectomy (2stS) — initially on the dominant hand, followed later by contralateral surgery. Only seven studies have directly studied these alternatives.

In 2013, Ibrahim et al.^17^ studied 270 patients, with 130 undergoing BTS and 140 undergoing 2stS. They found CS in 4.3% of the 2StS group, compared with 19.1% in the BTS group.

In 2015, Ravari et al.^18^ reported on 52 patients who underwent UniS, observing complete resolution of symptoms on the treated side in all patients (100%). They noted a 46.1% resolution contralaterally and found that 25% experienced CS. Only 7 patients (13.4%) sought contralateral surgery, and of those, 5 had CS.

Also in 2015, Youssef et al.^19^ analyzed 364 patients, reporting CS in 12.2% of the 2stS group versus 71.1% in the BTS group. No severe CS was noted in the 2stS group, and efficacy was reported at 96.7% for the 2stS compared to 97.2% for BTS, along with a satisfaction rate of 100% in the 2stS group versus 37.9% dissatisfaction in the BTS group.

In 2016, Menna et al.^20^ compared 135 patients with 2stS and 126 with BTS, with follow-up of more than 4 years, finding CS to be less frequent in the 2stS group (4.4% vs. 21.4%). They also reported greater QoL improvement in the 2stS group (≥ 95% vs. ≥ 90%), with statistical significance (P=0.001).

In 2022, Alkosha et al.^21^ compared 82 UniS patients with 112 BTS patients and found that CS was significantly lower in the UniS group; however, recurrence, satisfaction and QoL were similar after one year.

In 2023, Aikosha et al.^22^ used the same patient population and identified UniS as a negative predictor of CS. Their multivariate regression analysis indicated that BMI, plantar hyperhidrosis and unilateral VATS were significant predictors of CS.

Finally, in 2026, Shih et al.^23^ conducted the only prospective study with 118 patients (77 underwent BTS, and 41 UniS). They assessed patients pre– and 3-month post-operation, finding higher satisfaction in the UniS group (93% vs. 61%). Additionally, 80% of the UniS patients reported no CS, while 43% of the BTS group experienced mild CS (p=0.007). In the UniS group, 7.5% experienced improvement in the contralateral sweating, and 22% required contralateral sympathectomy.

Based on these studies, reducing HH on the dominant side, and potentially reducing it on the non-dominant side as well, is associated with a lower risk of CS. This reduction can lead to improved QoL and a decrease in the incidence and intensity of CS. However, there is still some uncertainty because the studies conducted lacked thorough evaluations, and only one prospective, multicenter, randomized trial has compared these two surgical strategies (unilateral versus one-stage bilateral) in terms of achieving HH resolution and preventing CS in a specific population (Japanese). This gap highlights the need for stronger evidence to determine the optimal surgical approach.

We conducted a randomized multicenter clinical trial across 11 institutions to test the hypothesis that unilateral thoracic sympathectomy would improve palmar HH while reducing the risk of CS compared with bilateral thoracic sympathectomy in a Brazilian population. The primary aim of the trial was to compare the reduction in hand sweating and the intensity of CS in participants with primary palmar HH who underwent one-stage bilateral thoracic sympathectomy versus those who underwent unilateral thoracic sympathectomy on the dominant side.

We also examined the number of patients who chose to undergo a contralateral sympathectomy, which is a two-stage bilateral procedure. We measured the incidence of CS in this group and compared it to both the unilateral and one-stage bilateral groups. Additionally, we assessed the postoperative QoL among participants who underwent BTS, 2stS, or UniS on the dominant side.

## 4. Patients and methods

### 4.1 Study design

This study was a prospective, randomized, controlled, multicenter clinical trial that enrolled 163 adult participants (age ≥18 years) with a body mass index ≤ 28 and a diagnosis of palmar HH. A total of 11 centers throughout Brazil participated in the study.^24^

All patients received similar treatment, following the same protocol, in accordance with the ethical standards of the Committee of Ethics for Analysis of Research Projects on Human Experimentation of the h and the principles outlined in the Declaration of Helsinki and is registered at Plataforma Brazil with CAAE 00273818.4.1001.0068 and has a ClinicalTrials.gov Identifier: NCT03921320. Written informed consent was obtained from every patient.

Subjects were randomly assigned to one of two treatment arms: the BTS Group, undergoing a one-stage bilateral thoracic sympathectomy (serving as the control group); or the UniS Group, receiving a unilateral thoracic sympathectomy on the dominant side (intervention arm). Patients in the UniS Group who were dissatisfied with their clinical outcome subsequently underwent a contralateral sympathectomy and constituted a third cohort — the two-stage bilateral group (2stS).

To randomize group assignment, we used a computer-generated random allocation sequence, and participants were randomly assigned in a 1:1 ratio to receive either UniS or BTS.

For the surgery, patients were given general anesthesia. The thermoablation segment was delineated between the upper edge of the 4th rib and the lower edge of the 5th costal arch, which corresponds to the location of the 4th sympathetic ganglion. In the bilateral group, the same procedure was also performed on the contralateral sympathetic chain. Starting from the 6th postoperative month, participants in the UniS group who felt the outcome unsatisfactory underwent surgery on the contralateral side, thus entering the 3rd group.

The patients in the UniS and BTS groups underwent clinical assessment, sweating intensity measurement, and QoL assessment at three time points: once before surgery and at 2 and 6 months after the operation. Patients in the 2ndS Group who underwent a second surgical procedure received two additional clinical assessments at 2 and 6 months following the second surgery.

The primary endpoints of the study were to assess the variation in sweating, as measured by the Hyperhidrosis HDSS^25^ and evaluate QoL using validated questionnaires (HidroQOl© and Horn)^26, 27^

administered at baseline and during follow-up visits.

The study aimed to determine sweating levels across all body regions and to assess QoL at the first visit before surgery. It also sought to examine improvements in sweating severity across all body regions, evaluate variations in QoL following surgery, and identify the presence or absence of CS.

To assess the intensity of sweating, we used the HDSS across 18 predefined body regions: right face, left face; right chest, left chest; right abdomen, left abdomen; right back, left back; right axilla, left axilla; right hand, left hand; right inguinal region, left inguinal region; right foot, left foot.

The HDSS is a validated tool for measuring HH severity, translated and validated into Portuguese for use across all body areas. This instrument consists of a single question that can be answered on a scale of four levels of sweat tolerance and its impact on the patient’s daily life. A Score of 1 indicates the absence of excessive sweating; a score of 2 indicates moderate HH, while scores 3 and 4 correspond to severe HH.

The palmar HH severity was defined as the sum of the values assigned to both hands in the HDSS.

The efficacy of sympathectomy was evaluated by the difference in the sum of values between the pre– and postoperative periods for both hands.

CS is defined as excessive sweating that develops after sympathectomy in body regions that did not previously exhibit clinically relevant sweating. To assess CS, we evaluated 16 body regions per patient that were non-sweating preoperatively (HDSS = 1) and followed their evolution over time. The outcome was defined by changes in HDSS: regions that remained at HDSS = 1 were classified as no CS; regions that increased to HDSS = 2 were classified as mild CS; and regions that increased to HDSS = 3 or 4 were classified as severe CS. This classification was applied separately to each assessed region. A patient was considered to have CS if at least one evaluated site developed CS.

To measure satisfaction, we used two instruments specifically developed for patients with HH: the Hyperhidrosis Quality of Life Index (HidroQOL) and the questionnaire described by Horn.

The HidroQOl is a validated questionnaire designed to assess everyday situations where HH may be a factor. It consists of an 18-question protocol divided into two domains: activities of daily living (6 questions) and psychosocial life (12 questions). Responses were scored on a 3-point scale (very = 2, a little = 1, no / not at all = 0). The total score could range from 0 to 36, with higher scores indicating greater impact on QoL levels:

The Horn questionnaire was developed in Brazilian Portuguese language and consists of 10 closed-ended questions. Each question offers four response options that indicate the impact on QoL: “nothing”, “a little”, “very”, and “very much”. The questionnaire scores range from 0 to 30.

The questions address embarrassment and the impact of symptoms on daily life, social events, physical activity, work, public speaking/meetings, perceived impression on others, self-esteem, leisure activities, the need for more frequent showers, and limitations in movement.

The total score could range from 0 to 30, with higher scores indicating greater impact on QoL levels.

### 4.2 Data analysis

#### Descriptive Statistical Analysis

Descriptive analyses were performed for all variables. Normally distributed continuous data were presented as mean ± standard deviation (SD), and non-normally distributed data as median (interquartile range [IQR], 25th–75th percentiles). Normality was assessed using the Shapiro–Wilk test (and visual inspection of histograms). Categorical variables were summarized as counts and percentages.

#### Inferential Statistical Analysis

For within-group comparisons of normally distributed variables, the paired Student’s t-test was used; for repeated measurements, repeated-measures analysis of variance (RM-ANOVA) was applied. For non-normally distributed variables, the Mann–Whitney U test and the Friedman test were used, as appropriate. Categorical variables were compared using the chi-square test, and Fisher’s exact test when expected cell counts were small. Correlations were assessed using Pearson’s correlation coefficient for normally distributed variables and Spearman’s rank correlation for non-normally distributed variables. All tests were two-sided with a type I error rate (α) of 0.05.

For the primary endpoint, between-group differences were evaluated using the unpaired Student’s t-test (or Mann–Whitney U test) and analysis of covariance (ANCOVA) adjusting for the corresponding baseline (pre-treatment) value. Unadjusted and adjusted means with 95% confidence intervals (95% CI) are reported. ANCOVA assumptions (homogeneity of regression slopes, equality of variances, and normality of residuals) were checked; when substantial violations were observed, an appropriate nonparametric alternative was used. In analyses involving non-randomized group comparisons, generalized linear models were applied to account for potential baseline imbalances between groups.

Correlation strength was interpreted as: very strong (|r| = 0.90–1.00), strong (|r| = 0.70–0.89), moderate (|r| = 0.40–0.69), weak (|r| = 0.20–0.39), and very weak (|r| = 0.00–0.19). All analyses were performed using SPSS version 21.0 (SPSS Inc., Chicago, IL, USA).

## 5. Results

A total of 163 patients were analyzed, and their baseline characteristics are summarized in Table 1. Most patients were women (62%) in the third decade of life (24.1 years) from the Heart Institute (26.4%). There was a wide distribution of HH types, with Palmar involvement present in all cases. The median BMI was 24.0 kg/m² with no significant difference between groups. Most patients were Caucasian (73.1%). Disease onset occurred in childhood (83.5%), with a median disease duration of 16 years.

**Table 1:** Baseline patient characteristics (pre-surgery) according to surgical group.

| Variable | TOTAL<br>(n = 163) | BTS<br>(n = 83) | UniS<br>(n = 80) | p-valor |
| --- | --- | --- | --- | --- |
| Age (years) | 24.1 [20.9; 30.5] | 23.3 [21.1; 28.5] | 26.0 [20.9; 31.7] | 0.179<sup>(a)</sup> |
| Gender |  |  |  | 0.106 |
| Male | 62 (38.0%) | 37 (44.6%) | 25 (31.2%) |  |
| Female | 101 (62.0%) | 46 (55.4%) | 55 (68.8%) |  |
| HH |  |  |  | **0.037** |
| Palmar | 24 (14.7%) | 10 (12.0%) | 14 (17.5%) |  |
| Palmar + Axillary | 7 (4.3%) | 7 (8.4%) | 0 (0.0%) |  |
| Palmar + Plantar | 91 (55.8%) | 48 (57.8%) | 43 (53.8%) |  |
| Palmar + Axillary + Plantar | 41 (25.2%) | 18 (21.7%) | 23 (28.8%) |  |
| BMI | 24.0 [21.0; 26.0] | 23.0 [20.5; 26.0] | 24.0 [21.0; 25.2] | > 0.999 |
| Ethnicity |  |  |  | 0.725 |
| Caucasian | 117 (73.1%) | 61 (74.4%) | 56 (71.8%) |  |
| African descent | 43 (26.9%) | 21 (25.6%) | 22 (28.2%) |  |
| Onset of illness |  |  |  |  |
| Childhood | 132 (83.5%) | 69 (84.1%) | 63 (82.9%) |  |
| Adolescence | 24 (15.2%) | 12 (14.6%) | 12 (15.8%) |  |
| Adult | 2 (1.3%) | 1 (1.2%) | 1 (1.3%) |  |
| Duration of illness (years) | 16.0 [12.2; 22.0] | 15.0 [11.0; 21.0] | 20.0 [13.5; 25.0] | 0.157<sup>(a)</sup> |

| Variable | TOTAL<br>(n = 163) | BTS<br>(n = 83) | UniS<br>(n = 80) | p-value |
| --- | --- | --- | --- | --- |
| HDSS dominant hand before surgery |  |  |  | 0.056 |
| Grade 1 | 1 (0.6%) | 1 (1.2%) | 0 (0.0%) |  |
| Grade 2 | 0 (0.0%) | 0 (0.0%) | 0 (0.0%) |  |
| Grade 3 | 16 (10.0%) | 4 (4.9%) | 12 (15.4%) |  |
| Grade 4 | 143 (89.4%) | 77 (93.9%) | 66 (84.6%) |  |
| HORN score before surgery | 20.3 ± 5.6 | 19.1 ± 6.5 | 19.7 ± 6.1 | 0.210 |
| HidroQOL before surgery | 27.0 [23.0; 31.0] | 28.0 [23.0; 31.0] | 27.0 [21.2; 30.0] | 0.386<sup>(a)</sup> |

(a) Non-parametric Wilcoxon-Mann-Whitney test
For the comparison of qualitative variables, Fisher's exact tests were performed for variables with two categories and Chi-square tests for the others.

Preoperatively, 89.4% had HDSS grade 4 (93.9% bilateral; 84.6% unilateral; p = 0.056). Mean preoperative HORN score was 20.3 ±5.6 (p = 0.210), and median HidroQoL was 27.0 (23.0–31.0), with no significant intergroup differences (p = 0.386), demonstrating that the patients suffered a lot in their hands, and the QoL in both groups was poor.

Table 2 summarizes HDSS responses for the right and left hands at baseline, 2 months, and 6 months in the BTS (n = 82) and UniS (n = 78) groups. Before surgery, nearly all patients in both groups had severe disease (HDSS 3–4) in both hands (BTS: 98.7%; UniS: 100%). At 2 months, >93% of BTS patients improved to HDSS 1–2 in both hands, whereas in the UniS group, 92.1% reached HDSS 1–2 in the operated (dominant) hand and 20.6% in the contralateral hand. At 6 months, 91.9% of BTS patients remained at HDSS 1–2 bilaterally; in the UniS group, 91.1% were HDSS 1–2 in the operated hand and 14.3% in the untreated hand. Overall, treated limbs showed marked improvement, and contralateral regression occurred in about one-seventh of unilateral cases (14.2%).

**Table 2:**
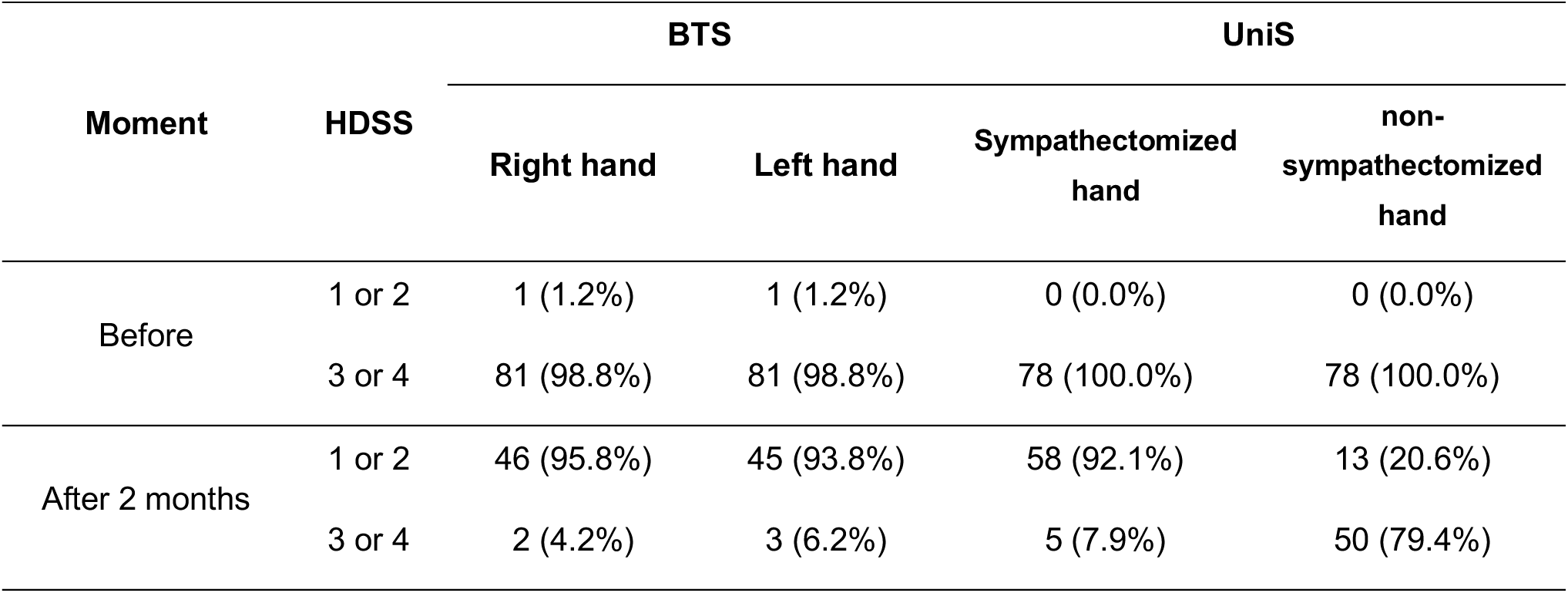
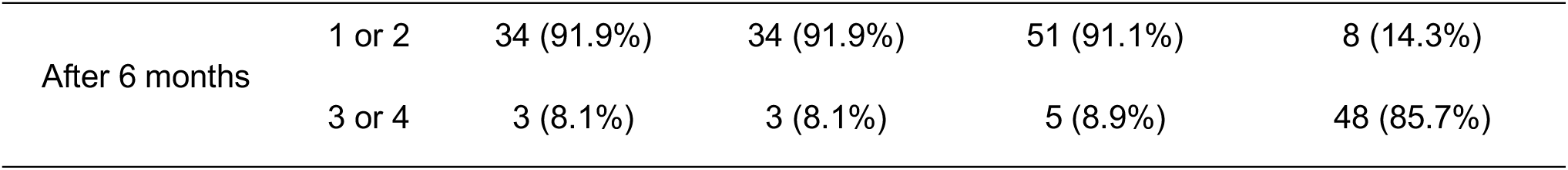
Evolution of HDSS hand scores in patients undergoing BTS vs. UniS sympathectomy.

Table 3 compares postoperative HDSS scores between the bilateral and unilateral groups. Preoperatively, both groups had a median HDSS of 4 in both hands. At 2 and 6 months, dominant-hand scores were similar between groups (median 1.0; p > 0.4), indicating comparable improvement. In contrast, for the non-dominant hand, the bilateral group maintained significantly lower scores at both time points (median 1.0), whereas the unilateral group remained higher (median 4.0 at 2 months and 3.0 at 6 months; p < 0.001).

**Table 3:** Hand Sweating Severity (HDSS) Scores Before Surgery and at 2 and 6 Months After Bilateral vs Unilateral Sympathectomy.

| Variável | BTS<br>(n = 82) | UniS<br>(n = 78) | p-valor |
| --- | --- | --- | --- |
| HDSS right hand before | 4.0 [4.0; 4.0] | 4.0 [4.0; 4.0] |  |
| HDSS right hand after 2 months | 1.0 [1.0; 1.0] | 1.0 [1.0; 2.0] | 0.482 |
| HDSS right hand after 6 months | 1.0 [1.0; 1.0] | 1.0 [1.0; 2.0] | 0.944 |
| HDSS left hand before | 4.0 [4.0; 4.0] | 4.0 [4.0; 4.0] |  |
| HDSS left hand after 2 months | 1.0 [1.0; 1.0] | 4.0 [3.0; 4.0] | **< 0.001** |
| HDSS left hand after 6 months | 1.0 [1.0; 1.0] | 3.0 [3.0; 4.0] | **< 0.001** |

The descriptive measures of the scores are presented as median and interquartile range [25; 75%].
The comparison tests were obtained from the adjustment of mixed models considering the appropriate distribution for each of the variables and including the interaction factor between group and time.

Table 4 summarizes the changes in right– and left-hand HDSS scores from baseline to the postoperative time points within each group. In both the bilateral and unilateral groups, the right hand showed a consistent 3-to 3-point reduction at 2 and 6 months compared with preoperative values (p < 0.001), indicating substantial symptom improvement. In the bilateral group, the left hand also improved significantly, with a −3 3-point reduction at both follow-ups. By contrast, in the unilateral group, the left hand showed no significant change at either postoperative assessment, with median changes of zero. Overall, bilateral sympathectomy improved symptoms in both hands, whereas unilateral sympathectomy provided sustained benefit mainly in the treated hand.

**Table 4:**
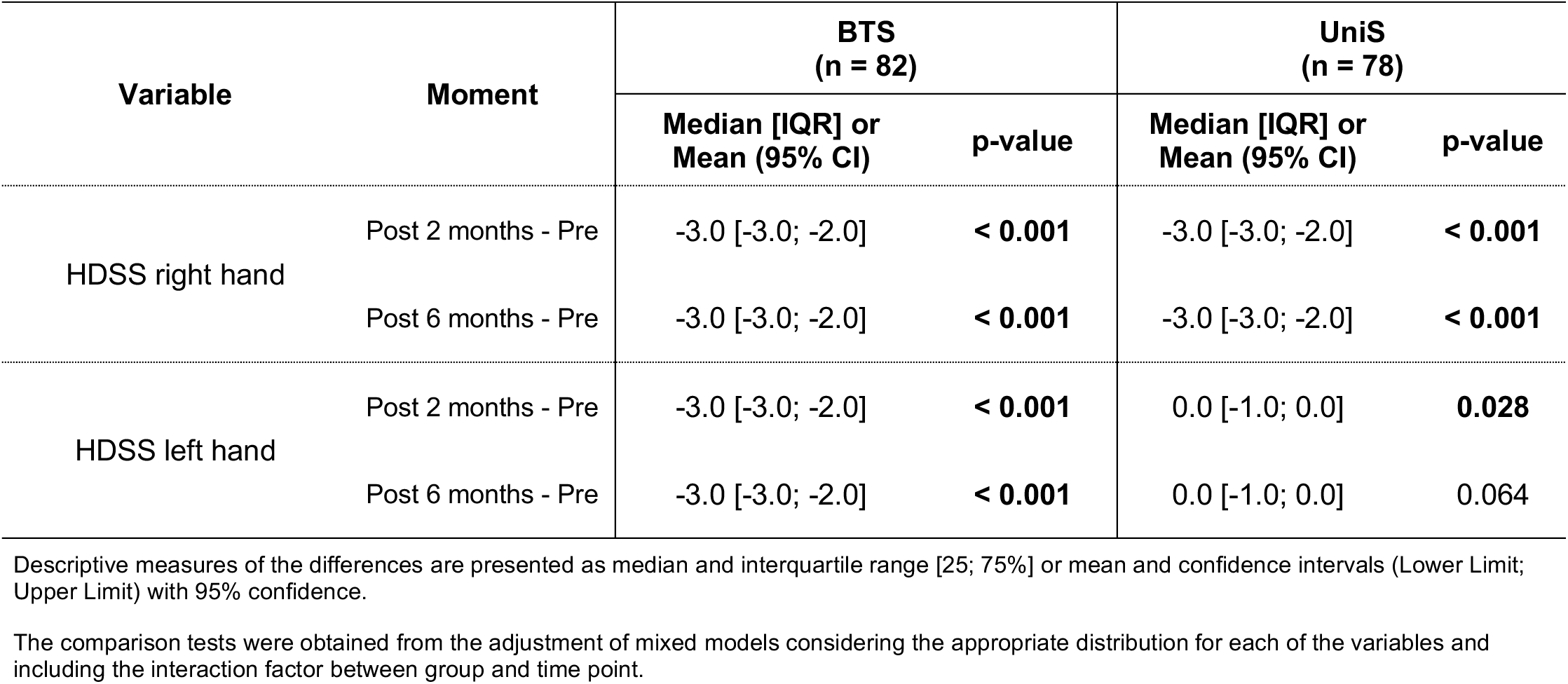
Comparison of the change in scores assessed between each of the post-surgery and pre-surgery time points, within each of the groups.

| Variable | Moment | BTS<br>(n = 82) |  | UniS<br>(n = 78) |  |
| --- | --- | --- | --- | --- | --- |
|  |  | Median [IQR] or<br>Mean (95% CI) | p-value | Median [IQR] or<br>Mean (95% CI) | p-value |
| HDSS right hand | Post 2 months - Pre | -3.0 [-3.0; -2.0] | **< 0.001** | -3.0 [-3.0; -2.0] | **< 0.001** |
|  | Post 6 months - Pre | -3.0 [-3.0; -2.0] | **< 0.001** | -3.0 [-3.0; -2.0] | **< 0.001** |
| HDSS left hand | Post 2 months - Pre | -3.0 [-3.0; -2.0] | **< 0.001** | 0.0 [-1.0; 0.0] | **0.028** |
|  | Post 6 months - Pre | -3.0 [-3.0; -2.0] | **< 0.001** | 0.0 [-1.0; 0.0] | 0.064 |

Descriptive measures of the differences are presented as median and interquartile range [25; 75%] or mean and confidence intervals (Lower Limit; Upper Limit) with 95% confidence.
The comparison tests were obtained from the adjustment of mixed models considering the appropriate distribution for each of the variables and including the interaction factor between group and time point.

Table 5 compares the severity of compensatory sweating (CS) at 6 months after surgery between the BTS (n = 47) and UniS (n = 62) groups. The proportion of patients without CS was virtually identical across groups, affecting about one quarter of participants in each group. The unilateral group showed a higher rate of mild CS (53.2% vs 34.1%), whereas the bilateral group showed a higher rate of severe CS (40.4% vs 21.0%). Thus, despite similar rates of absent CS, CS tended to be more severe after BTS than after the UniS.

**Table 5:** Distribution of compensatory sweating grades 6 months after surgery between the bilateral and unilateral groups.

| CS | BTS<br>(n = 47) | UniS<br>(n = 62) | p-value<br>Fisher test |
| --- | --- | --- | --- |
| absent | 12 (25.5%) | 16 (25.8%) |  |
| light | 16 (34.1%) | 33 (53.2%) |  |
| severe | 19 (40.4%) | 13 (21.0%) | 0,0344 |

Table 6 compares postoperative Horn and HydroQOL scores between the bilateral and unilateral groups. Preoperatively, scores were comparable. At 2 months, the bilateral group had markedly lower median Horn (1.5 vs. 8.0) and HydroQOL (6.0 vs. 18.0) scores than the unilateral group (p < 0.001), indicating a stronger early response. This difference persisted at 6 months, with lower Horn (5.0 vs. 12.0) and HydroQOL (8.0 vs. 17.0) scores in the bilateral group (p < 0.001), suggesting sustained benefit. Overall, the larger reductions in scores with bilateral sympathectomy reflect greater clinical improvement and quality-of-life gains, whereas the unilateral group maintained higher scores over time.

**Table 6:** Comparison of scores assessed between the groups at post-surgery time points.

| Variable | BTS<br>(n = 82) | UniS<br>(n = 78) | p-value |
| --- | --- | --- | --- |
| Horn score before | 21.0 [16.0; 25.0] | 20.0 [15.0; 24.0] |  |
| Horn score after 2 months | 1.5 [0.0; 5.0] | 8.0 [4.0; 15.5] | **< 0.001** |
| Horn score after 6 months | 5.0 [1.0; 9.0] | 12.0 [4.0; 16.0] | **< 0.001** |
| HidroQOL before | 28.0 [23.0; 31.0] | 27.0 [21.2; 30.0] |  |
| HidroQOL after 2 months | 6.0 [3.0; 15.0] | 18.0 [11.5; 23.0] | **< 0.001** |
| HidroQOL after 6 months | 8.0 [5.0; 14.0] | 17.0 [10.8; 25.0] | **< 0.001** |

Descriptive measures of scores are presented as median and interquartile range [25; 75%].
The comparison tests were obtained from the adjustment of mixed models considering the appropriate distribution for each of the variables and including the interaction factor between group and time point.

Table 7 summarizes pre– to postoperative changes in Horn and HydroQOL scores for the bilateral and unilateral groups. In both arms, Horn scores declined significantly at 2 and 6 months versus baseline, indicating symptom improvement; however, the magnitude of change was greater in the bilateral group. HydroQOL scores also decreased significantly at both follow-ups in both groups, consistent with better quality of life, with more extensive reductions after bilateral sympathectomy. All comparisons were statistically significant (p < 0.001). This consistent separation between groups suggests that while either strategy is beneficial relative to baseline, bilateral surgery yields a more robust overall response across both instruments and time points.

**Table 7:**
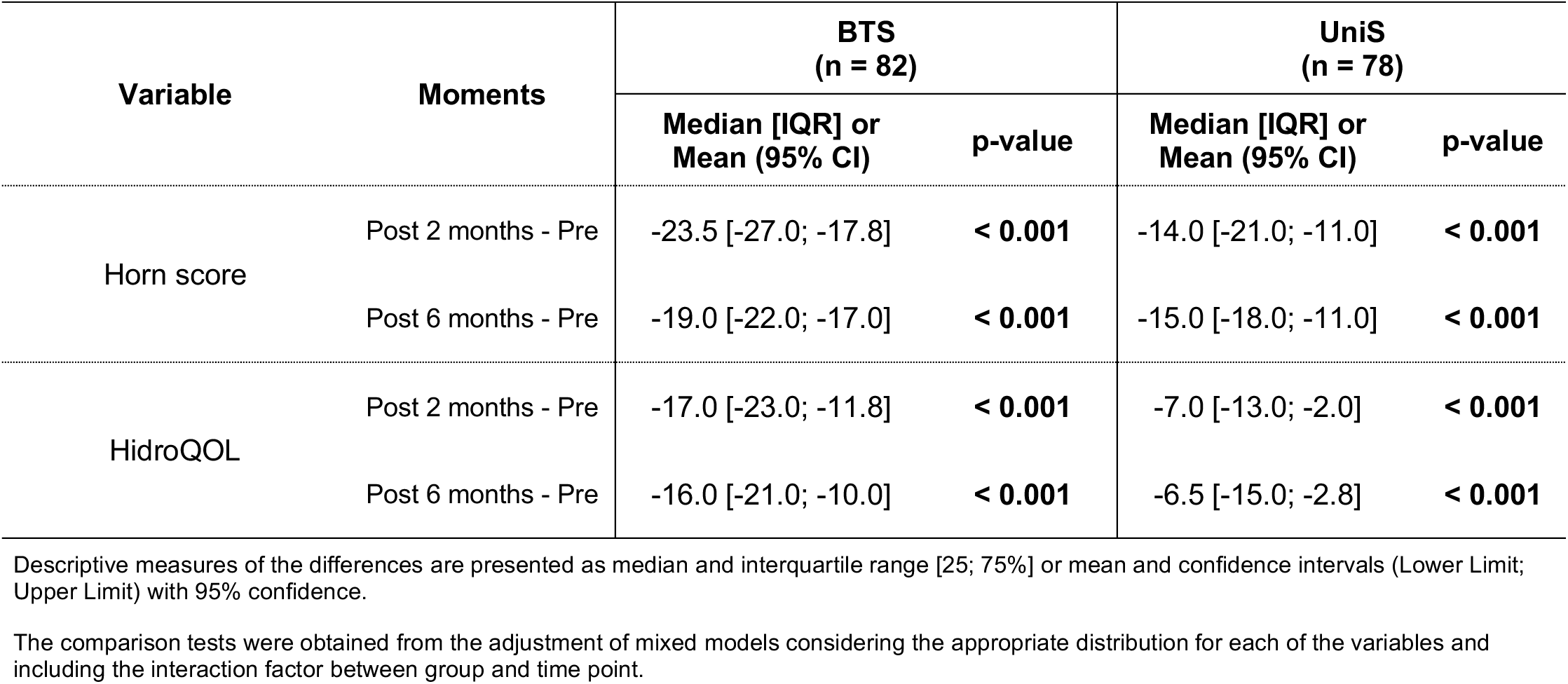
Comparação da alteração dos escores avaliados entre cada um dos momentos pós e o momento pré-cirurgia, dentro de cada um dos grupos.

| Variable | Moments | BTS<br>(n = 82) |  | UniS<br>(n = 78) |  |
| --- | --- | --- | --- | --- | --- |
|  |  | Median [IQR] or<br>Mean (95% CI) | p-value | Median [IQR] or<br>Mean (95% CI) | p-value |
| Horn score | Post 2 months - Pre | -23.5 [-27.0; -17.8] | < 0.001 | -14.0 [-21.0; -11.0] | < 0.001 |
|  | Post 6 months - Pre | -19.0 [-22.0; -17.0] | < 0.001 | -15.0 [-18.0; -11.0] | < 0.001 |
| HidroQOL | Post 2 months - Pre | -17.0 [-23.0; -11.8] | < 0.001 | -7.0 [-13.0; -2.0] | < 0.001 |
|  | Post 6 months - Pre | -16.0 [-21.0; -10.0] | < 0.001 | -6.5 [-15.0; -2.8] | < 0.001 |

Descriptive measures of the differences are presented as median and interquartile range [25; 75%] or mean and confidence intervals (Lower Limit; Upper Limit) with 95% confidence.
The comparison tests were obtained from the adjustment of mixed models considering the appropriate distribution for each of the variables and including the interaction factor between group and time point.

After the initial procedure, 16 patients (25.8%) elected contralateral surgery on the non-dominant limb, and all showed improvement in sweating severity. Among those with preoperative HDSS 4 (n = 13), postoperative scores improved to grade 1 in 8 patients, grade 2 in 2, and grade 3 in 1. Among patients with preoperative HDSS 3 (n = 4), three improved to grade 1 and one to grade 2. One patient with preoperative HDSS 2 improved to grade 1; notably, this patient requested the second procedure primarily because of severe contralateral axillary symptoms (HDSS 4). The HDSS is a 4-point, patient-reported severity scale commonly used in hyperhidrosis research. Compensatory sweating—an established adverse effect after sympathectomy—occurred in 9 of 16 patients (56.2%): seven had mild CS at a single site, one had mild CS at two sites, and one developed intense CS (a 2-point increase in two assessed sites).

Table 8 tracks HDSS, Horn, and HidroQOL trajectories in unilateral patients who later underwent contralateral surgery. The dominant hand (operated first) showed an early and sustained HDSS reduction, already significant at 2 months and continuing to improve at 6, 8, and 12 months (all p < 0.001). In contrast, the contralateral hand showed no significant change at 2 and 6 months, but improved sharply after the second procedure, with a 3-point decrease in HDSS at 8 and 12 months (p < 0.001). Horn scores declined progressively over follow-up, with larger drops after contralateral surgery, exceeding 24-point reductions at 8 and 12 months. HidroQOL followed the same pattern—modest early improvement, then a marked, significant decrease after contralateral surgery at 8 and 12 months. Overall, these findings suggest the staged bilateral strategy yields more robust and consistent clinical and quality-of-life gains.

**Table 8:**
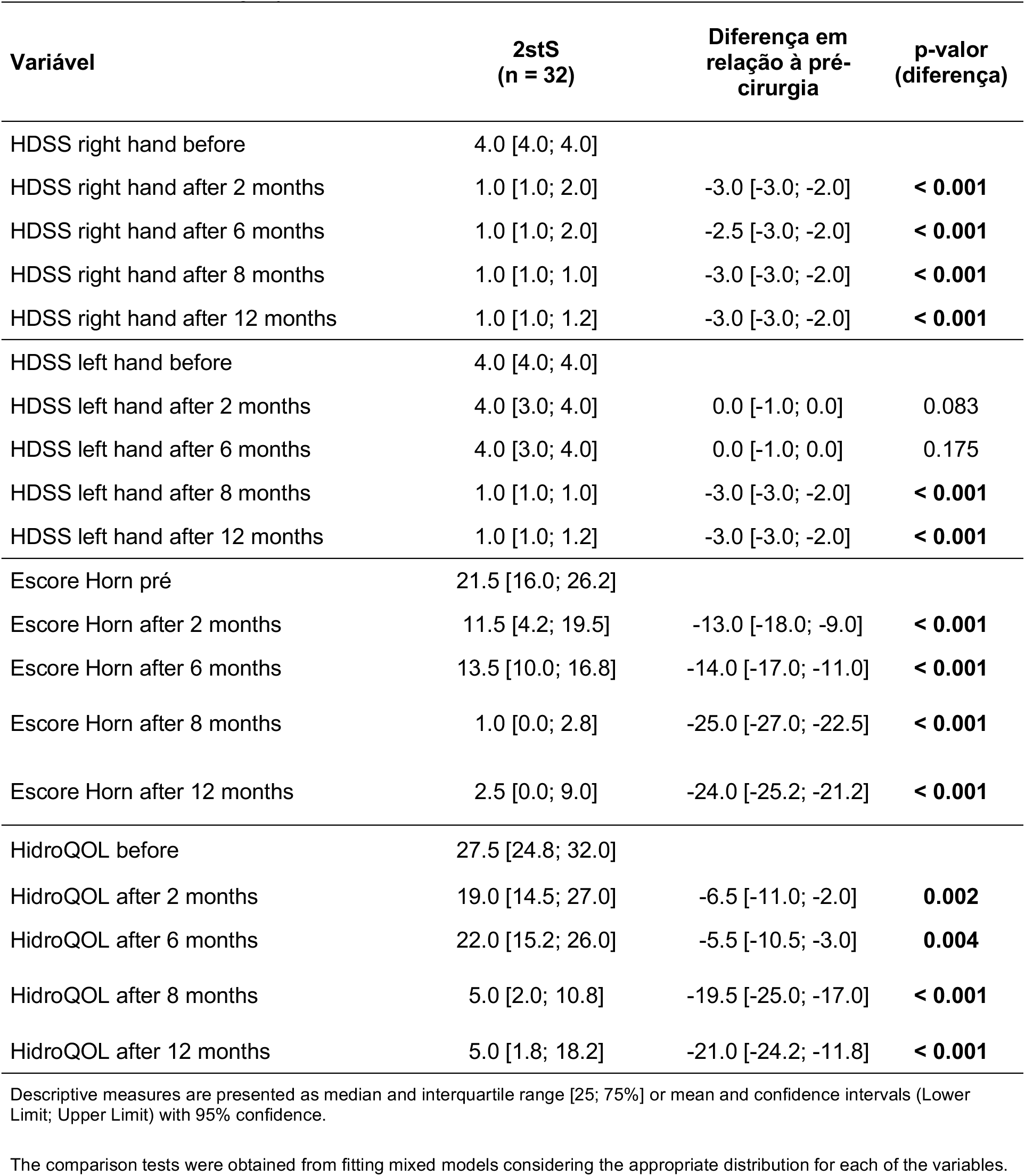
Comparison of scores assessed for patients in the 2stS surgery between each of the post– and pre-surgery time points.

| Variável | 2stS<br>(n = 32) | Diferença em<br>relação à pré-cirurgia | p-valor<br>(diferença) |
| --- | --- | --- | --- |
| HDSS right hand before | 4.0 [4.0; 4.0] |  |  |
| HDSS right hand after 2 months | 1.0 [1.0; 2.0] | -3.0 [-3.0; -2.0] | **< 0.001** |
| HDSS right hand after 6 months | 1.0 [1.0; 2.0] | -2.5 [-3.0; -2.0] | **< 0.001** |
| HDSS right hand after 8 months | 1.0 [1.0; 1.0] | -3.0 [-3.0; -2.0] | **< 0.001** |
| HDSS right hand after 12 months | 1.0 [1.0; 1.2] | -3.0 [-3.0; -2.0] | **< 0.001** |
| HDSS left hand before | 4.0 [4.0; 4.0] |  |  |
| HDSS left hand after 2 months | 4.0 [3.0; 4.0] | 0.0 [-1.0; 0.0] | 0.083 |
| HDSS left hand after 6 months | 4.0 [3.0; 4.0] | 0.0 [-1.0; 0.0] | 0.175 |
| HDSS left hand after 8 months | 1.0 [1.0; 1.0] | -3.0 [-3.0; -2.0] | **< 0.001** |
| HDSS left hand after 12 months | 1.0 [1.0; 1.2] | -3.0 [-3.0; -2.0] | **< 0.001** |
| Escore Horn pré | 21.5 [16.0; 26.2] |  |  |
| Escore Horn after 2 months | 11.5 [4.2; 19.5] | -13.0 [-18.0; -9.0] | **< 0.001** |
| Escore Horn after 6 months | 13.5 [10.0; 16.8] | -14.0 [-17.0; -11.0] | **< 0.001** |
| Escore Horn after 8 months | 1.0 [0.0; 2.8] | -25.0 [-27.0; -22.5] | **< 0.001** |
| Escore Horn after 12 months | 2.5 [0.0; 9.0] | -24.0 [-25.2; -21.2] | **< 0.001** |
| HidroQOL before | 27.5 [24.8; 32.0] |  |  |
| HidroQOL after 2 months | 19.0 [14.5; 27.0] | -6.5 [-11.0; -2.0] | **0.002** |
| HidroQOL after 6 months | 22.0 [15.2; 26.0] | -5.5 [-10.5; -3.0] | **0.004** |
| HidroQOL after 8 months | 5.0 [2.0; 10.8] | -19.5 [-25.0; -17.0] | **< 0.001** |
| HidroQOL after 12 months | 5.0 [1.8; 18.2] | -21.0 [-24.2; -11.8] | **< 0.001** |

Descriptive measures are presented as median and interquartile range [25; 75%] or mean and confidence intervals (Lower Limit; Upper Limit) with 95% confidence.
The comparison tests were obtained from fitting mixed models considering the appropriate distribution for each of the variables.

## 6. Discussion

Excessive palmar sweating, which is the definition of PH, can cause substantial impairment in daily life, leading to professional limitations, social embarrassment, and difficulties in affective and interpersonal relationships. In more severe cases, PH may contribute to avoidance behaviors and even social phobia.^11, 28^

Nowadays, first-line treatment consists of oxybutynin hydrochloride, which improves symptoms in approximately 70% of patients, regardless of baseline disease characteristics.^29, 30, 31^ When medical therapy fails or is not tolerated, surgery becomes the most effective option. However, sympathectomy may be followed by compensatory sweating (CS), which can negatively impact satisfaction.^32^ To mitigate this risk, in this study, we explored the staged approach: unilateral sympathectomy on the dominant side followed by contralateral sympathectomy only if the patient considers the residual symptoms unacceptable after the initial procedure.

This study design was valuable because it tested two different strategies addressing our main hypothesis for reducing CS after sympathectomy: a) that UniS is feasible, practical, acceptable and clinically useful; and b) that staged bilateral Surgery may lead to less CS than a single-session bilateral procedure. Moreover, in implementing a multicentric trial, we avoided a purely explanatory or efficacy trial design. Efficacy trials often include small, highly selected samples from specialized centers, potentially overestimating benefits and underestimating harms. In contrast, current guidance emphasizes real-world effectiveness in broader populations. Our trial aligns with a pragmatic approach: recruitment, intervention delivery, follow-up, and outcome assessment are consistent with routine practice. Using the PRECIS-2 (Pragmatic-Explanatory Continuum Indicator Summary) framework, the present study can be classified as pragmatic, aiming to generate evidence that supports adoption into everyday care.^33, 34^

We analyzed 163 patients with palmar hyperhidrosis who underwent sympathectomy, comparing BTS (n=83) and UniS (n=80). Baseline characteristics were comparable between groups: most participants were Caucasian individuals (73.1%), female (62%), young (median 24.1 years), with a median BMI of 24.0 kg/m², with disease onset predominantly having occurred in childhood (83.5%), and with prolonged disease duration (median 16 years). These characteristics are consistent with most studies published in Brazil and worldwide.^35,36^

The broad geographical distribution of participating centers confers ethnic and climatic diversity, which is relevant for an event motivated by these factors. Given Brazil’s continental dimensions and regional variations, the 3,876 km distance between the northern and southern centers reinforces the external validity of the study. It reduces the likelihood that local particularities significantly impacted the results.

To ensure a robust and standardized outcome assessment, we used validated instruments to quantify both sweating severity and quality of life. Sweating was measured with the HDSS across 18 body sites at every follow-up, enabling a detailed evaluation of anatomical distribution patterns. Quality of life was assessed with two widely validated questionnaires (HidroQOL and Horn), integrating internationally recognized patient-reported outcomes with clinical severity metrics. In this multicenter, multiracial cohort, this structured approach strengthens the reliability of our findings by combining objective grading with comprehensive, validated QoL measures.^25, 26, 27^

In our study, all patients underwent T4 sympathectomy, a resection level supported by evidence suggesting effective control of palmar hyperhidrosis with lower compensatory seating rates.^37^ As expected in surgical candidates, most patients presented with severe symptoms preoperatively (HDSS 3–4), followed by a rapid and sustained improvement after surgery, with HDSS decreasing to 1–2 in most cases. In the BTS group, over 90% of patients improved to HDSS 1–2 in both hands at 2 and 6 months, consistent with large series reporting >90% clinical improvement in target areas (palmar/axillary), with excellent immediate control and maintenance during follow-up.^38^

When BTS was compared with UniS, the operated hand in the UniS group showed outcomes similar to the BTS (92% in HDSS 1–2 at 2 months and 91% at 6 months), suggesting comparable local efficacy on the operated side. These findings have already been described in studies exploring unilateral, bilateral or staged approaches, in which the benefit in the treated hand is consistently high. In contrast, the untreated hand in the UniS group remained predominantly severe (HDSS 3–4) in most patients (79% at 2 months and 86% at 6 months).^7, 17, 18, 20, 21^ However, 14% of patients showed improvement in the contralateral hand (untreated) without the need for immediate sympathectomy, indicating that for individuals with bilateral involvement, the unilateral approach controls the contralateral hand in approximately 1 in 7 patients. This “cross-effect” may reflect central autonomic modulation, individual variability, or the limitations of semi-quantitative instruments such as the HDSS. In summary, our findings corroborate that in the Brazilian population, unilateral sympathectomy offers excellent control in the treated limb and contralateral benefit in 1/7 of cases after 6 months.

The postoperative onset of sweating in previously unaffected body regions is termed CH. It is a well-recognized yet incompletely understood phenomenon after thoracic sympathectomy. Although its pathophysiology remains uncertain, CH is commonly hypothesized to reflect a compensatory thermoregulatory response mediated by central autonomic mechanisms. In published series, CH most often involves the back and abdomen and is the most frequently reported complication of video-thoracoscopic sympathectomy (VTS).^39^ In our study, 75% of patients in both groups reported CH at some point after surgery. Severe CH, defined as an increase in HDSS from 1 to 3–4 at any anatomical site, occurred at 6 months in 40.4% of the BTS group and 21.0% of the UniS group, a statistically significant difference. Severe CH may substantially impair quality of life and can generate symptoms comparable in burden to those reported preoperatively, albeit in different body regions, contributing to postoperative dissatisfaction.^40^

We acknowledge that the relatively high incidence of compensatory hyperhidrosis (CH) may partly reflect our tropical setting, where higher temperatures can increase sweating; nevertheless, CH can occur across climates.^41^ Importantly, CH was assessed systematically. All patients underwent detailed preoperative mapping of sweating sites, and postoperative changes were quantified with site-specific HDSS scoring. CH was defined as new onset sweating in previously unaffected areas (preoperative HDSS = 1), and severity was graded by the magnitude of HDSS increase.^42^

Because CH risk correlates with the extent and level of sympathetic interruption, it is notable that both groups underwent the same resection level (T4). With this factor controlled, our data suggest that interrupting the chain only on the dominant side—while preserving the contralateral chain—may reduce severe CH, consistent with prior reports and supported here by a standardized assessment approach.^43, 44^

Several studies indicate that sympathectomy can improve quality of life even when complications such as compensatory sweating (CS) occur, because relief of palmar symptoms and functional limitations often outweighs adverse effects.^3, 6, 9, 23, 24, 33^ In our study, we used two validated instruments that capture complementary domains of hyperhidrosis burden: the Horn Score (social and emotional impact) and HidroQOL (physical, psychological, and functional domains).

Preoperatively, both groups reported similarly poor quality of life, with no differences in Horn (21.0 vs. 20.0) or HidroQOL (28.0 vs. 27.0), confirming a high baseline burden. At 2 months, quality of life improved in both groups. Still, gains were greater after bilateral sympathectomy, with larger reductions in Horn (1.5 vs. 8.0) and HidroQOL (6.0 vs. 18.0). This likely reflects more complete sweating control, translating into meaningful benefits in daily activities, work, and social interactions. The pattern persisted at 6 months, with the bilateral group maintaining lower scores (Horn: 5.0 vs. 12.0; HidroQOL: 8.0 vs. 17.0; p < 0.001), indicating sustained benefit.

Overall, bilateral surgery yielded a more robust and durable improvement in QoL, while unilateral surgery still produced substantial gains versus baseline; the higher residual scores likely reflect persistent sweating on the untreated side, especially during bimanual tasks. Concordant findings across both instruments support the link between greater symptom control and greater functional and psychosocial improvement, and mixed-model analyses with group-by-time interaction reinforced internal validity.^40^

Only 18 patients (29.0%) from the group initially treated with unilateral sympathectomy chose to undergo contralateral surgery. This low crossover rate suggests that, for most patients, the clinical improvement and quality-of-life gains achieved after the first intervention were sufficient, particularly given the option to complete surgery on the opposite side at any time.

One illustrative case involved a patient whose contralateral hand was only mildly affected at baseline (HDSS grade 2) and improved to grade 1 without contralateral surgery. The indication for a second procedure in this patient was driven instead by persistent severe axillary sweating (HDSS = 4) on the untreated side. This example highlights that decisions about staged completion are often guided by individualized residual symptoms rather than hand sweating alone and underscores the importance of reassessing the full symptom profile during follow-up.

Among all patients who underwent contralateral sympathectomy, we observed an apparent, clinically meaningful reduction in sweating on the newly operated side, while maintaining the sustained anhidrosis achieved on the previously treated dominant limb. These findings indicate that a two-stage approach preserves the benefit of the initial operation and remains consistently effective when the second procedure is performed later. Overall, contralateral surgery provides additional symptom control when needed, with a direct and relevant impact on postoperative severity in the treated regions.^17, 18, 19^

After contralateral surgery, the incidence of compensatory sweating (CS) was 56.2%. Most patients developed mild CS limited to a single anatomical site; only one patient reported mild CS at two sites, and just one case was classified as intense CS involving two evaluated sites. This distribution suggests that, although CS is relatively frequent after staged completion, its severity in this setting is generally mild to moderate. These findings are consistent with previous reports in the literature and support the notion that severe CS is uncommon when contralateral sympathectomy is performed as a second-stage procedure.^17,18, 19^

Given its prevalence and potential impact on satisfaction, CS should be addressed explicitly during the informed consent process, particularly when discussing bilateral strategies performed in two stages. Clear preoperative counseling can help align expectations, facilitate shared decision-making, and contextualize the trade-off between improved symptom control and the risk of new sweating in other body regions.

Quality-of-life outcomes, assessed with the Horn Score and HidroQOL, corroborated the clinical improvement observed throughout follow-up. Both instruments showed progressive improvement over time, with the most pronounced reductions occurring after contralateral surgery. Horn Score decreases exceeded 24 points, and HidroQOL also improved significantly at 8 and 12 months, indicating that quality-of-life gains were not only early but sustained. These trajectories were consistent with the HDSS findings, reflecting symptom reduction and fewer functional and psychosocial limitations.

Taken together, our results suggest that a staged bilateral approach can deliver clinical and quality-of-life benefits comparable to those achieved with primary bilateral procedures, while maintaining an acceptable complication profile. For patients with hyperhidrosis, this strategy offers an effective surgical option that enables stepwise symptom control and substantial quality-of-life improvement, while allowing sequential, individualized management of the expected treatment-related consequences.

The study has limitations of a circumstantial, methodological, and design nature. The main limitation was that part of the study was conducted during the COVID pandemic. This severely limited the inclusion of new participants and considerably hampered the follow-up of patients who had already undergone surgery. It is well known that during the pandemic, outpatient clinics for benign diseases in most public hospitals were temporarily closed.^42^

Another important limitation is the use of self-report measures of disability rather than objective functional or biological assessments. Accurate quantitative measurements of sweating are not feasible because sweating flow is not constant throughout the day, and no instrument is sufficiently practical to be incorporated into clinical routine. Consequently, outcomes are assessed through indirect instruments such as HDSS and QoL questionnaires. This means that the results derive from responses that depend on subjective assessment and the participant’s memory.

Finally, another study limitation was the inclusion of participants with palmar and axillary hyperhidrosis. Some of these participants, after unilateral sympathectomy, chose to proceed to the contralateral surgery not because of discomfort from sweating in the non-dominant hand, but because of armpit sweating. In other words, the existence of axillary sweating contaminated the results regarding palmar sweating.

## 7. Conclusions

In this multicenter Brazilian study, bilateral sympathectomy provided an immediate and substantial reduction in sweating in both upper limbs, with a marked improvement in quality of life, albeit at the cost of higher rates of compensatory sweating (CS). Unilateral sympathectomy also produced a robust decrease in sweating in most treated limbs. It yielded a contralateral benefit in approximately 1 in 7 patients, translating into quality-of-life improvement, though generally to a lesser extent than with the bilateral approach.

When implemented as a staged, bilaterally performed strategy (initially treating the dominant limb, followed by contralateral sympathectomy only if needed), this pathway can ultimately deliver clinical and quality-of-life outcomes comparable to those of primary bilateral surgery. Importantly, it also allows a subset of patients to avoid a second procedure altogether, either because they experience bilateral improvement after unilateral surgery or because the benefit achieved on the dominant side is sufficient to meet their expectations. For those who proceed to completion, outcomes appear similar to those achieved with one-stage bilateral sympathectomy.

These findings should be clearly discussed with patients to support shared decision-making and individualized surgical planning. By presenting the trade-offs among unilateral, bilateral, and staged bilateral strategies, clinicians can help patients more accurately weigh functional benefits, the risk of CS, and expected long-term satisfaction when choosing the approach that best fits their priorities and symptom profile.

## Data Availability

All data produced in the present study are available upon reasonable request to the authors

